# Placental growth plays a key role in the link between maternal glucose levels in pregnancy and risk of preeclampsia

**DOI:** 10.1101/2025.10.23.25338694

**Authors:** Annika Jaitner, Robin N. Beaumont, Alice E. Hughes, Krasimira Tsaneva-Atanasova, Jack Bowden, Rachel M. Freathy

## Abstract

Preeclampsia is a serious condition affecting 2-4% of pregnancies globally. However, it is more common in pregnancies with diabetes (e.g. 10-20% in Type 1 diabetes) than in pregnancies without diabetes. In the general population, higher maternal fasting glucose levels are observationally associated with higher risk of preeclampsia, but whether the relationship is causal is unknown. Based on previous research, we hypothesise that higher fasting glucose increases placental weight, likely through fetal insulin mediated growth, and higher placental weight increases the risk of preeclampsia.

We used data from published genome-wide association studies on fasting glucose (N=281,416), placental weight (Nfetal=65,405, Nmaternal=61,228) and preeclampsia (Nfetal=377,975, Nmaternal=723,181) and individual level data from the Exeter Family Study of Childhood Health (N=948) and the Avon Longitudinal Study of Parents and Children (N=5,214). We compared observational multivariable regression analysis with Mendelian Randomization (MR), an analytical approach that uses genetic variants to estimate causal effects of exposures (e.g. fasting glucose) on outcomes (e.g. risk of preeclampsia).

We found that genetically instrumenting a 1mmol/l higher fasting glucose caused a higher placental weight of 44.16g (95%CI: [29.82,58.49]). Furthermore, we showed that fetal genetic predisposition to higher placental weight increased the risk of preeclampsia (OR=1.99, 95% CI: [1.25, 3.19]). Using the two-step MR method, we estimated that a maternal genetic predisposition to higher fasting glucose increases the risk of preeclampsia through increasing placental weight with an OR of 1.26 (95% CI: [1.05,1.49]).

We found evidence for a causal effect of higher maternal fasting glucose levels on risk of preeclampsia, and our results are consistent with this effect being mediated via placental weight. Further well-powered studies are needed to confirm the causal relationship between higher maternal fasting glucose and preeclampsia. If confirmed, our findings suggest research focusing on fetal insulin may help elucidate preeclampsia disease mechanisms.

**Research in context:** *Evidence before this study:* Observational analysis from the Hyperglycaemia and Adverse Pregnancy Outcome (HAPO) study showed that 1 standard deviation higher maternal fasting glucose was associated with a 1.21 (95% confidence interval: [1.13-1.29]) higher odds of preeclampsia. However, whether this relationship is causal is unknown. It is known that maternal glucose crosses the placenta, but maternal insulin does not. The glucose reaching the fetus, stimulates fetal insulin secretion which acts as a potent growth factor for both the fetus and placenta. Moreover, insights from genetic analyses recently revealed that a higher placental weight is causally linked to higher preeclampsia risk.

*Added value of this study:* Building on the above findings, we hypothesised that higher maternal glucose results in increased fetal insulin-mediated placental growth that increases the risk of preeclampsia. To our knowledge, this is the first study to investigate the relationships between maternal fasting glucose, placental weight and preeclampsia in the same study. The use of genetic data allows us to compare observational findings with causal analyses. Our results provide evidence that greater placental growth is on the causal pathway between higher maternal fasting glucose levels in pregnancy and risk of preeclampsia. Therefore, we highlight a potential additional contributor to the diabetes-preeclampsia link: fetal insulin mediated growth of the placenta.

*Implications of all the available evidence:* Our findings contribute to an understanding of why preeclampsia is more prevalent in pregnancies affected by diabetes. A better understanding of causal biological processes underlying the pathology of preeclampsia is important for the development of treatment and optimal care for pregnant women. If confirmed, our results support that measuring and optimising maternal glucose levels in pregnancies will be an important tool in reducing preeclampsia risk and suggest that markers of placental growth could help with risk stratification in the future.

## Introduction

Preeclampsia is a serious condition, affecting 2-4% of pregnancies globally ^1^. It is a major cause of maternal and fetal morbidity and mortality ^2^. Preeclampsia is diagnosed by the onset of hypertension after 20 weeks of gestation and at least one other complication, including proteinuria, acute kidney or liver injury, and uteroplacental dysfunction ^3^. Time of diagnosis classifies preeclampsia into either early (≤34 weeks of gestation) or late onset (> 34 weeks of gestation)^3^. To date, early onset preeclampsia is better understood than late onset preeclampsia and believed to arise from abnormal placentation ^3^. While there is a higher per-pregnancy risk of complications in early onset preeclampsia, late onset, also referred to as “term” preeclampsia, is three times more common and still carries a substantial risk of adverse pregnancy outcomes ^4^.

Preeclampsia is more common in mothers with diabetes; it is diagnosed in 15-20 % and in 10-14 % of mothers with type 1 and type 2 diabetes respectively ^5^. Preeclampsia is also more common in mothers with gestational diabetes mellitus. Catalano et al. (2012) ^6^ showed an incidence of 5.9% and 3.5% of preeclampsia cases in non-obese mothers (body mass index at 28 weeks of gestation < 33 kg/m^2^) with and without gestational diabetes mellitus, respectively. Yang et al. (2022) ^2^ highlight in their review that several population-based studies have shown that gestational diabetes mellitus is associated with the occurrence of preeclampsia. Some of those studies conclude that obesity is the main contributing factor, since higher BMI is a key risk factor for both GDM and higher blood pressure, a key maternal risk factor for preeclampsia. While common risk factors like maternal age and higher maternal body mass index could explain some of the association between preeclampsia and type 2 or gestational diabetes, type 1 diabetes is an autoimmune disease not generally related to body habitus and older age and has the highest frequency of preeclampsia amongst diabetes pregnancies. Therefore, the underlying mechanisms to explain the higher rate of preeclampsia in pregnancies complicated by diabetes are not fully understood.

Maternal glucose levels, which are higher in mothers with diabetes, could play a key role. Observational analysis from the Hyperglycaemia and Adverse Pregnancy Outcome (HAPO) study (not stratified by gestational diabetes mellitus or obesity status) showed that a 0.4 mmol/l (1 standard deviation (SD)) higher maternal fasting glucose level was associated with a 1.21 (95% confidence interval (CI): [1.13-1.29]) higher odds of preeclampsia ^7^. While higher maternal glucose levels could directly affect placental function, it is possible that the fetal response plays a role in the aetiology of preeclampsia. Mother’s glucose crosses the placenta, but maternal insulin does not ^8^. The glucose reaching the fetus, stimulates fetal insulin secretion ^8^. Fetal insulin is a known growth factor important for fetal and placental growth. Hughes et al. (2023) ^9^ showed that fetal insulin accounts for approximately half of human birth weight at term. Furthermore, genetic studies of common and rare variants have shown that fetal insulin is an important placental growth factor ^10,11^.

Placental growth in late pregnancy is implicated in preeclampsia pathophysiology. Placental function reduces as the placenta outgrows the uterine capacity: the chorionic villi become more closely packed, restricting the intervillous space and resulting in syncytiotrophoblast stress ^12^. The importance of high fetoplacental demand in preeclampsia is illustrated by higher prevalence of preeclampsia in multiple pregnancies ^13^. Moreover, in a recent study, Beaumont et al. (2023) ^10^ showed, using genetic analyses, that a higher placental weight is causally linked to higher preeclampsia risk.

Building on this knowledge, we hypothesise that higher maternal glucose increases fetal insulin secretion, resulting in placental growth that increases the risk of preeclampsia (figure 1). It is challenging to test this hypothesis and tease apart cause and effect in the context of pregnancy. However, the availability of large-scale genetic datasets offers an opportunity to understand causal relationships. Fetal and maternal genotypes may be used in a Mendelian randomization (MR) approach to test for potential causal associations between maternal or fetal traits and pregnancy outcomes. MR is an effective method analogous to a randomized controlled trial: genotypes, which are randomly allocated at conception, are independent of factors that normally confound a conventional epidemiological study ^14^. They can therefore be used to proxy exposures and estimate possible causal associations with outcomes. In this paper we combine multivariable regression and different Mendelian randomization techniques to investigate the relationship between fasting glucose, placental weight and preeclampsia risk using individual level data from two pregnancy cohorts and data from five large genome-wide association studies (GWAS).

**Figure 1.**
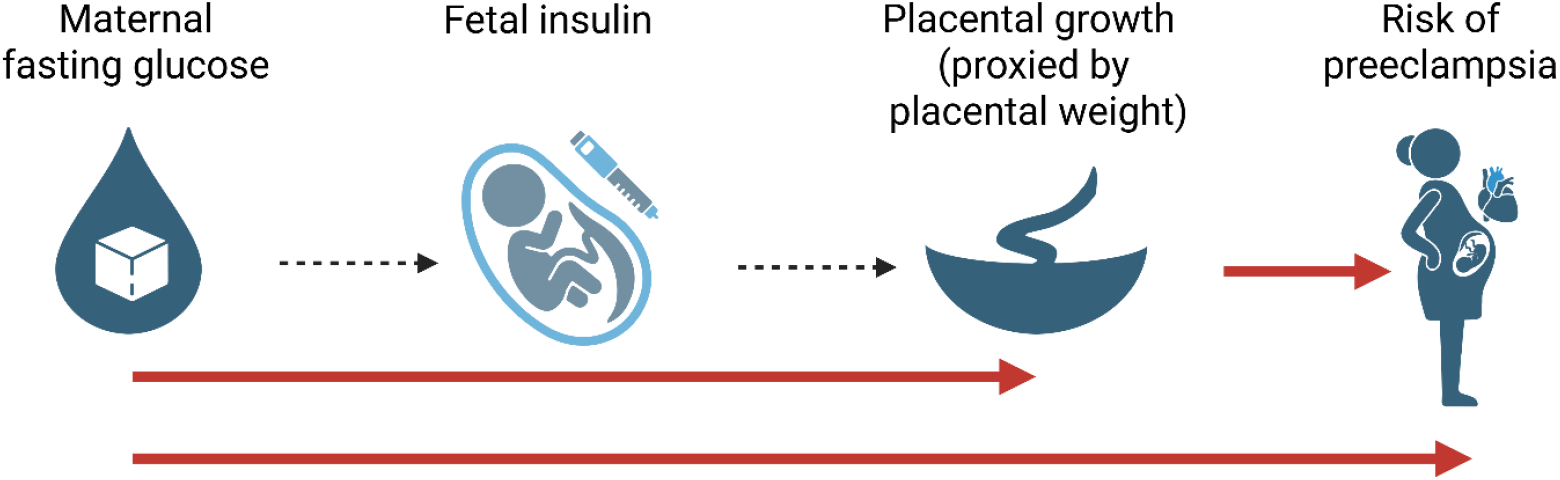
Biological hypothesis of the relationship between maternal glucose and risk of preeclampsia. Red arrows highlight the pathways we have tested and described in detail in this paper. Created with BioRender.com.

## Methods

### Individual level data from different cohorts

We used individual level data from the **Exeter Family Study of Childhood Health (EFSOCH)** and the **Avon Longitudinal Study of Parents and Children (ALSPAC)**. We only included singleton pregnancies with a gestational duration greater or equal to 37 weeks. Full details of the studies, a flowchart of further exclusions and descriptive statistics of the cohorts are given in the Supplementary material (figure S1, table S1-S2).

### Summary data from GWAS

Additionally, we used data on single nucleotide polymorphisms (SNPs) shown to be associated with fasting glucose (measured in plasma), placental weight and preeclampsia in large GWAS. Details on the GWAS studies are given in the Supplementary Material (table S3). For placental weight and preeclampsia SNPs, we used effect estimates that had been adjusted for the correlation between maternal and fetal genotypes (r ∼ 0.5) so that they would indicate maternal genetic effects conditional on fetal genotype, and vice versa. The adjustment was performed using a weighted linear model and is explained in more detail elsewhere ^15^.

### Multivariable regression analysis

We performed linear and logistic regressions using individual level data from EFSOCH and ALSPAC to assess the observational relationships between fasting glucose, placental weight and preeclampsia. All observational analyses were adjusted for offspring birth weight, offspring sex, gestational duration, maternal age, parity and whether the mother smoked at any point in pregnancy. The unadjusted estimates can be found in the Supplementary Material (table S4). We also performed the regression analysis without adjustment for birth weight and gestational duration as those two phenotypes could act as colliders for fasting glucose, placental weight and preeclampsia (table S5). Additionally, we compared the observational association between maternal fasting glucose and risk of preeclampsia with estimates previously reported by the HAPO study ^7^, which was conducted in 15 centres in 9 countries and yielded a total sample size of 25,505 individuals including 4.8 % of preeclampsia cases. Their analysis adjusted for field centre, age, body mass index, height, smoking, alcohol use, family history of diabetes, gestational age at oral glucose tolerance test, baby’s sex, parity, and family history of hypertension and prenatal urinary tract infection. It is important to note that the HAPO study results were not restricted to term born babies only.

### Mendelian randomization (MR)

Mendelian randomization (MR) utilises genetic variants as instrumental variables to proxy a modifiable exposure, to estimate its causal effect on a health outcome. More details including relevant assumptions are given in the Supplementary material.

#### One-sample MR

One-sample MR was performed using individual level data from EFSOCH and ALSPAC. We used genetic variants that have been shown to associate with the exposures of interest (fasting glucose and placental weight) in the respective GWAS described in table S3. Due to risk of weak instrument bias, especially in the small individual level data studies available to us, we combined the genetic instruments into a single, strong instrument instead of using the SNPs individually. We calculated a weighted genetic risk score (GRS) for individuals using the effect sizes from the respective external GWAS as weights. We then performed two-stage least square (2SLS) analyses, regressing the exposure on the GRS and then subsequently, the outcome on the genetically predicted exposure. Both stages of the 2SLS were adjusted for the first 10 ancestry principal components (PCs) and either the maternal genotype (if a fetal GRS was used in the first stage) or fetal genotype (if a maternal GRS was used in the first stage). Additionally, the analysis in EFSOCH was adjusted for genotyping batch. The regression model is given in the Supplementary material. The estimation of the standard error for the causal estimate, obtained in the second stage of the analysis, accounted for first-stage uncertainties.

#### Two-sample MR

For two-sample MR analysis, summary statistics from GWAS were used. We took the independent SNPs that showed an association with our exposures of interest based on the results of the respective GWAS. Next, those SNPs and their effect sizes were extracted from the outcome GWAS. If SNPs were not present in the outcome GWAS, they could not be included in the analysis. Harmonising the effect sizes for the instruments ensured that the effect of the SNP corresponded to the same allele in the exposure and the outcome GWAS data sets. The number of SNPs used for each two-sample MR analysis and F-statistics to assess instrument strength are given in table S6. Generally, a F-statistic above 10 is recommended ^14^. We report the inverse variance weighted two-sample MR analyses as our main result. For all analyses the TwoSampleMR R-package was used ^16^.

#### Two-step MR

To assess the effect of fasting glucose on risk of preeclampsia via placental weight we performed two-step MR. Here the effect of maternal fasting glucose on placental weight is multiplied by the effect of placental weight on the risk of preeclampsia. Those effects were obtained from the two-sample MR analyses. The standard error for the two-step MR estimate was calculated with the delta method (formula given in the Supplementary Material) ^17^.

### Effective sample size

For multivariable regression and one-sample MR analyses with preeclampsia as the outcome, we calculated an effective sample size, which represents the sample size for an equivalently powered study within a balanced sample of 50% controls and 50% cases ^18^. The formula for the calculation is given in the Supplementary Material.

### Sensitivity analyses

We investigated violation of the MR assumptions by performing sensitivity analyses for the two-sample MR analyses. To account for potential pleiotropy, we performed MR-Egger ^19^, MR-GRIP ^20^, weighted median MR ^21^ and weighted mode MR analyses ^22^. We used the Q-statistic to assess potential heterogeneity. We then performed MR-Radial to identify and remove potential outliers, thereby reducing the heterogeneity ^23^.

### Validation in pregnancy of fasting glucose SNPs from non-pregnant cohorts

For our two-sample MR analyses, we used the biggest GWAS for fasting glucose to date ^24^ to obtain the strongest instrument. However, the 105 SNPs and effect sizes were identified in non-pregnancy cohorts. We refer to those as “all fasting glucose SNPs”. To validate our assumption that those SNPs are relevant in pregnancy, we calculate a weighted GRS and tested its association with maternal fasting glucose measured at 28 weeks of gestation in EFSOCH (figure S2). For additional validation, we perform our two-sample MR analyses with fasting glucose as the exposure with another set of SNPs taken from a GWAS performed in women of Chinese ancestry with fasting glucose levels in pregnancy ^25^. We refer to those as “during pregnancy SNPs (Chinese)”.

## Results

### Higher maternal fasting glucose increases placental weight

The adjusted observational analysis in EFSOCH (N = 816) showed a 23.55 g (95% CI: [3.15, 43.94]) higher placental weight per 1 mmol/l higher fasting glucose.

The one-sample MR analysis yielded a causal effect estimate of 210.87 g (95% CI: [92.35, 329.39]) higher placental weight per 1 mmol/l higher genetically predicted maternal fasting glucose. This analysis was performed in 545 individuals in EFSOCH and the F-statistic for this analysis was 40.82. Beaumont et al. 2023 ^10^ (using 33 fasting glucose SNPs from Scott et al (2012) ^26^ showed previously that genetically instrumenting a 1mmol/l higher fasting glucose caused a 47.40g (95% CI: [23.70, 71.20]) higher placental weight using inverse variance weighted two-sample MR analysis. We repeated the two-sample MR analysis using a larger number of fasting glucose SNPs from a more recent GWAS: we used 99 SNPs from the fasting glucose GWAS by Chen et al. 2021 ^24^ (6 removed SNPs due to unavailability in the placental weight GWAS are given in table S6). The F-statistic for this analysis was 94.18. In line with the previously published results, we found that genetically instrumenting a 1mmol/l higher fasting glucose caused a higher placental weight of 44.16g (95%CI: [29.82,58.49]). The “during pregnancy SNPs (Chinese)” results are discussed in the sensitivity analysis section. All results are summarised in figure 2.

**Figure 2.**
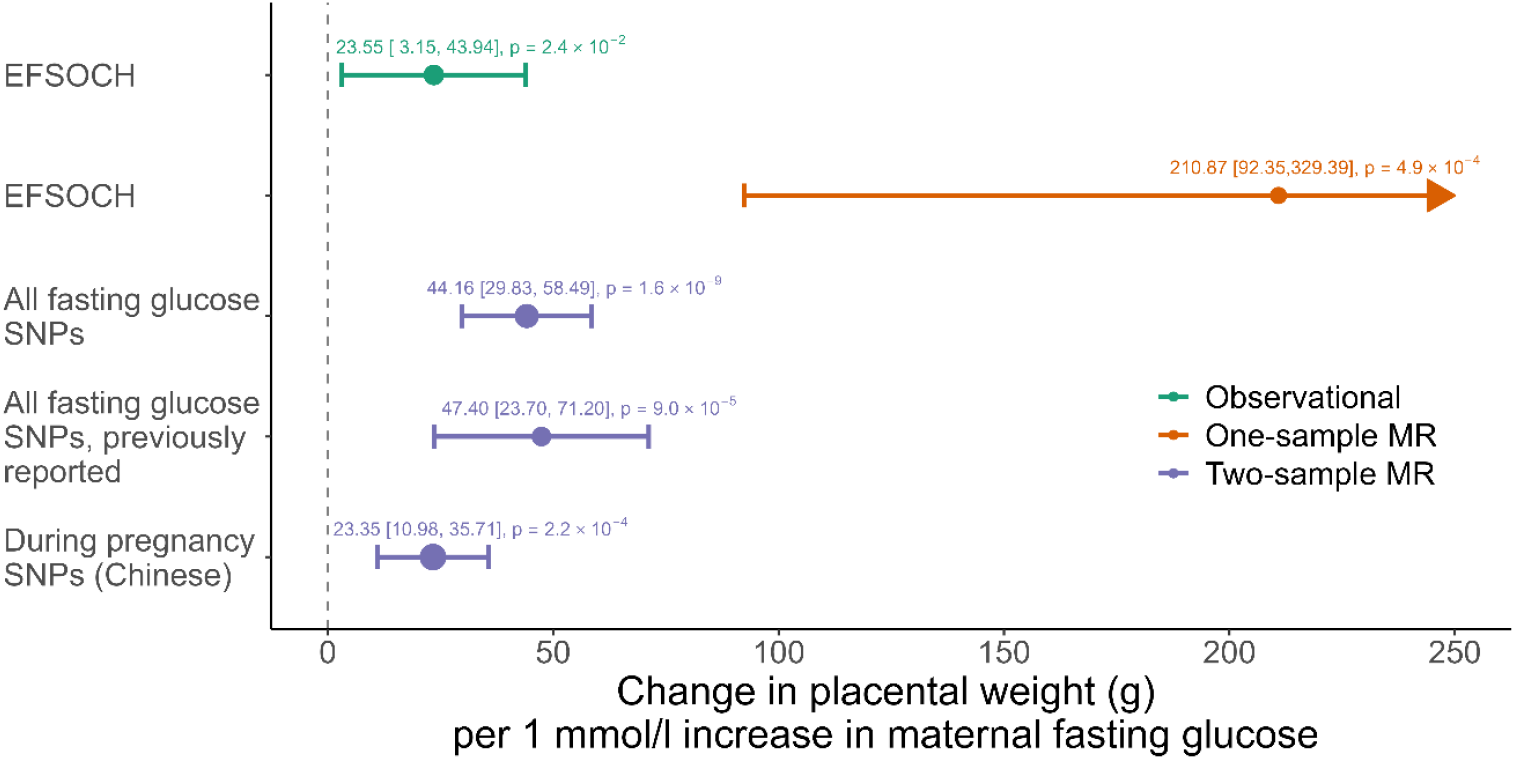
Observational and causal effects of maternal fasting glucose on placental weight using individual and summary level data. The previously reported result is taken from Beaumont et al. 2023 ^10^. 95% CI are shown in squared brackets.

### Higher placental weight increases the risk of preeclampsia

Our analysis did not support a non-zero observational association between higher placental weight and increased risk of preeclampsia in EFSOCH (OR = 0.90, 95% CI: [0.53, 1.52] per 1SD (132.5g) increase in placental weight) nor ALSPAC (OR = 1.12, 95% CI: [0.84, 1.51] per 1SD increase in placental weight), performed in N = 813 (effective sample size 89) and N = 4,831(effective sample size 299) respectively.

We performed the one-sample MR analysis in ALSPAC using a weighted fetal GRS to genetically predict placental weight. Those results indicated a positive causal effect of genetically predicted higher placental weight on increased risk of preeclampsia (OR = 10.40, 95% CI: [1.84, 58.75]). The analysis was performed in 2,045 individuals, which included 31 preeclampsia cases. The effective sample size is therefore 122. The F-statistic for this analysis was 34.08. The sample size for the same analysis in EFSOCH was much smaller with 548 (14 preeclampsia cases, effective sample size 55) individuals. Therefore, the results for this analysis are too imprecise to be meaningful and are not shown.

We performed two different two-sample MR analyses using either maternal or fetal SNP effects associated with placental weight. We showed that fetal genetic predisposition to higher placental weight increased the risk of preeclampsia (OR = 1.99, 95% CI: [1.25, 3.19]). For this analysis 34 SNPs were used resulting in an F-statistic of 24.31. The results aligned with the findings from Beaumont et al. 2023 ^10^(OR = 1.82, 95% CI: [1.23, 2.69]), which used the same SNPs associated with placental weight but a different outcome GWAS as the biggest maternal GWAS for preeclampsia ^27^ was not available at the time of their analysis. The maternal GWAS for preeclampsia is necessary for the WLM adjustment of the fetal effect sizes. We also estimated that maternal genetic predisposition to higher placental weight increases the risk of preeclampsia with an OR of 3.67 (95% CI: [1.76, 7.62], F-statistic: 28.97). Note that this analysis was performed using only 3 SNPs (rs2168101, rs180435, rs303998). All results are summarised in figure 3.

**Figure 3.**
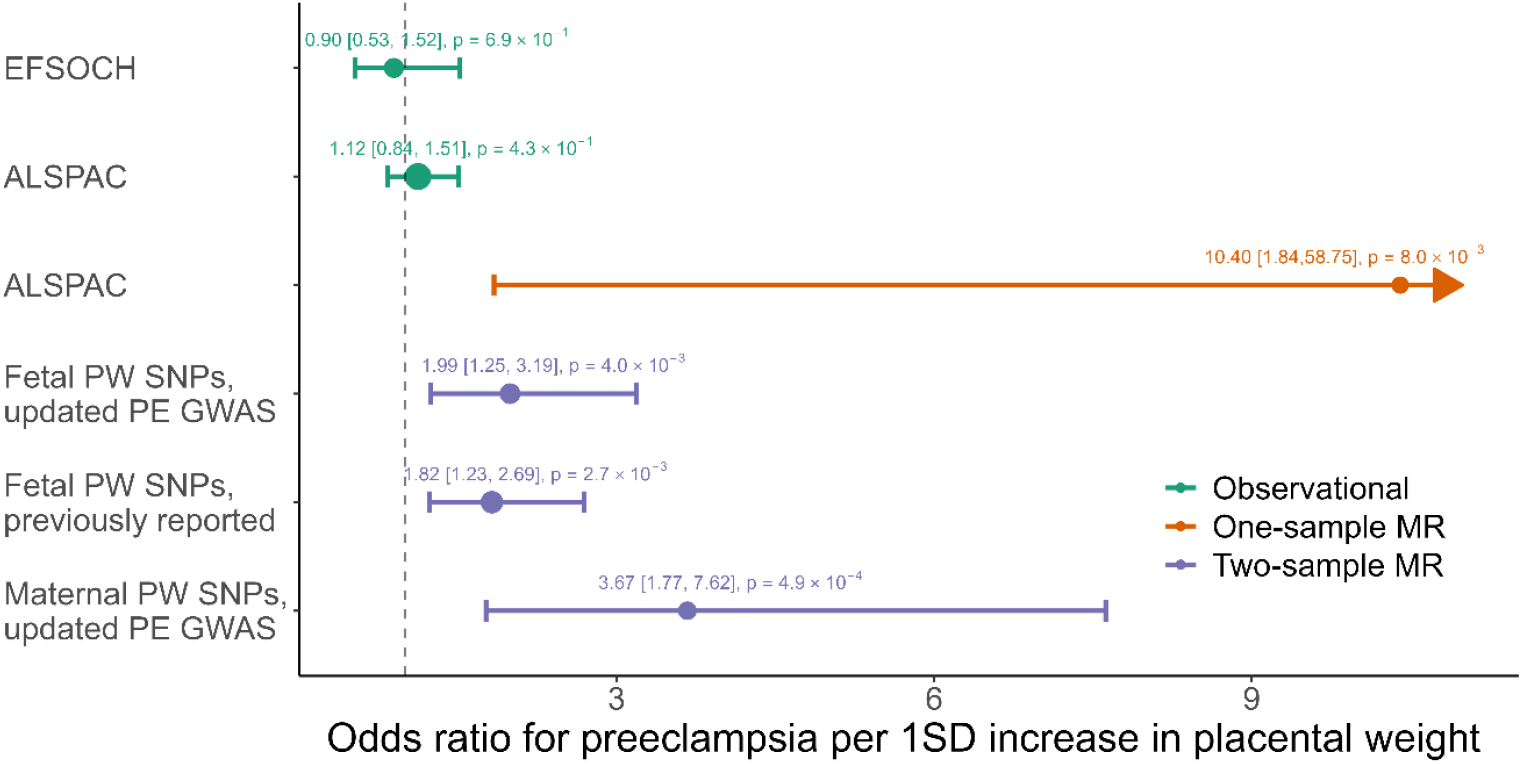
Observational and causal effects of placental weight on risk of preeclampsia using individual and summary level data. Previously reported results were taken from Beaumont et al. 2023 ^10^. PW: Placental weight, PE: Preeclampsia. 95% CI are shown in squared brackets.

### Evidence of a causal effect of maternal fasting glucose on risk of preeclampsia

The observational association of maternal fasting glucose with risk of preeclampsia obtained from the EFSOCH study (OR = 4.02, 95% CI: [1.63, 9.91]) aligned with what has been previously reported by the HAPO study (OR = 1.62, 95% CI: [1.36, 1.90]). The confidence intervals for our analysis in EFSOCH are wide, likely due to the small sample size of 896 individuals with 25 preeclampsia cases. One-sample MR analysis for this pathway was not conclusive due to the small number of individuals available in EFSOCH for this analysis (effective sample size 59).

Our two-sample MR analysis directionally aligned with the observational association but had wide 95% confidence intervals crossing the null (OR = 1.26 (95% CI: [0.85, 1.86], F-statistic: 95.25) per genetically predicted 1mmol/l increase in fasting glucose).

Using the two-step MR method, we estimated that a maternal genetic predisposition to higher fasting glucose increases the risk of preeclampsia through increasing placental weight with an OR of 1.26 (95% CI: [1.05,1.49]). The results are summarised in figure 4.

**Figure 4.**
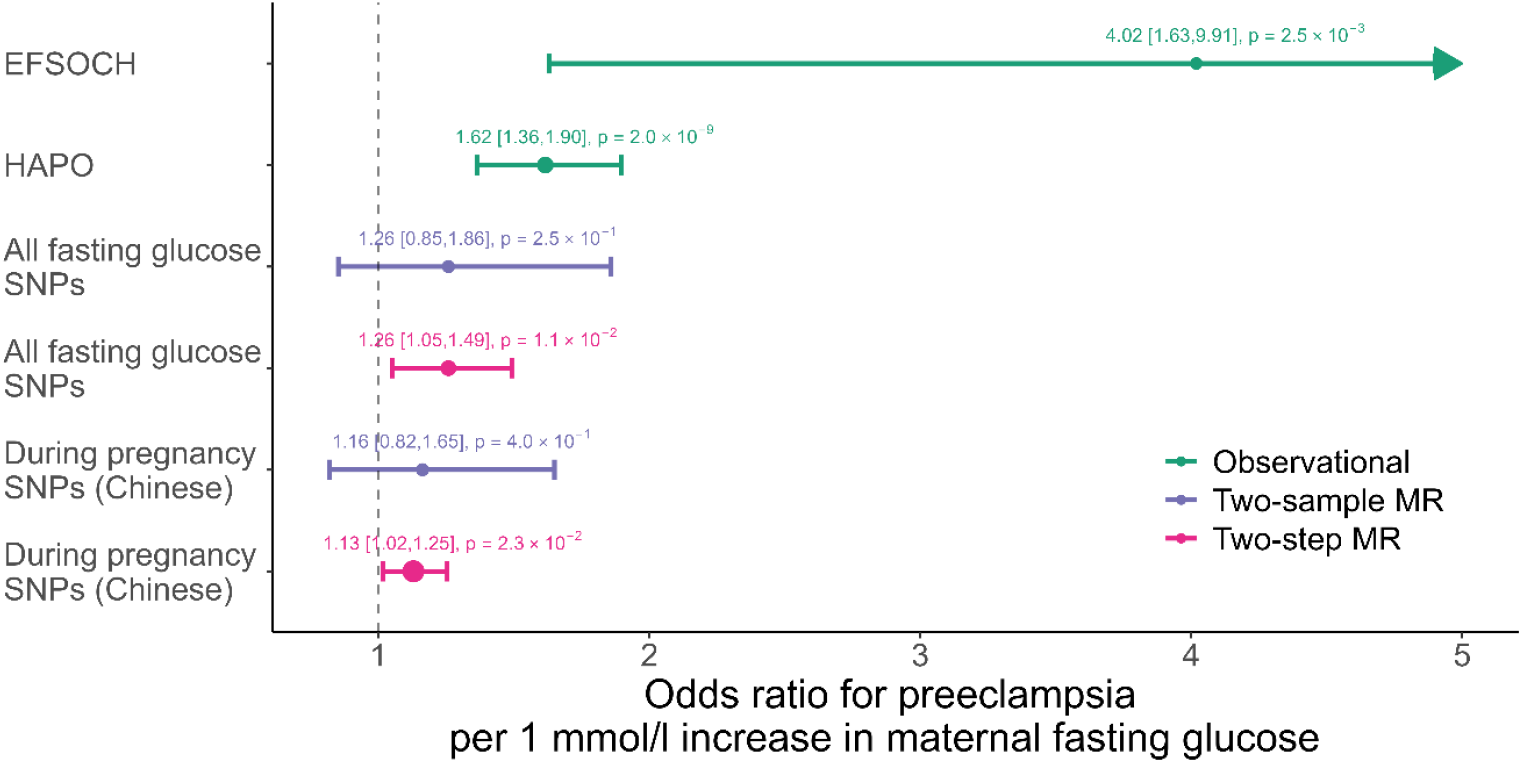
Observational and causal effects of maternal fasting glucose on risk of preeclampsia using individual and summary level data. The result for the HAPO study was taken from their own publication ^7^. 95% CI are shown in squared brackets.

### Sensitivity analysis

The unadjusted observational associations in EFSOCH and in ALSPAC for all three pathways as well as the regression results adjusted for offspring sex, maternal age, parity and smoking status (but not for birth weight and gestational duration) were consistent with the results shown in figure 2-4. While the magnitude of the estimates changed depending on the adjustment, the direction of the effect was consistent and did not change the interpretation of our results (table S4-S5).

The weighted genetic risk scores generated in EFSOCH using “all fasting glucose SNPs” ^24^ and “during pregnancy SNPs (Chinese)” ^25^ both showed an association with maternal fasting glucose measured at 28 weeks of gestation. The association was stronger for the “all fasting glucose SNPs” weighted genetic risk score (figure S2). The strong, positive associations of both set of SNPs with pregnancy fasting glucose in the EFSOCH cohort means that they are suitable to genetically instrument fasting glucose in pregnancy. Using “during pregnancy SNPs (Chinese)” yielded a causal effect of 23.35g (95% CI: [10.98, 35.71], F-statistic: 107.93) higher placental weight per 1 mmol/l increase in fasting glucose, consistent with our main result. The estimated effect of fasting glucose on risk of preeclampsia was also consistent with the main two-sample MR results (OR = 1.16, 95% CI: [0.82, 1.65], F-statistic: 106.88).

Similarly, the two-step MR analyses using the “during pregnancy SNPs (Chinese)” is in line with the result using “all fasting glucose SNPs” (OR = 1.13, 95% CI: [1.02,1.25]).

The various two-sample MR sensitivity analyses remained consistent with the inverse variance weighted two-sample MR results (figures S3-S5). Removing outliers using MR-radial improved the heterogeneity statistic and in some cases the F-statistic (table S7). The two-sample MR results after outlier removal were similar to the results before outlier removal (figures S3-S5) and aligned with our interpretation of the results.

## Discussion

Using individual level data from two cohorts and results from five large GWAS, we have shown that higher maternal fasting glucose increases the risk of preeclampsia likely via an increase in placental weight. While some of these pathways have been explored in different studies previously, to our knowledge, this is the first study to investigate the relationships between maternal fasting glucose, placental weight and preeclampsia in the same study. Our findings contribute to an understanding of why preeclampsia is more prevalent in mothers with diabetes. While previous studies have identified common risk factors for preeclampsia and gestational diabetes ^2,3^, those risk factors are often not present in mothers with type 1 diabetes, who themselves have the highest rates of preeclampsia. Our work highlights a potential additional contributor: fetal insulin-mediated growth of the placenta.

Our results from multivariable linear regression, one-sample and two-sample MR showed that higher fasting glucose increases placental weight. These results are in line with the biological hypothesis that higher fetal insulin secretion in response to higher maternal glucose levels increase placental growth. Shields et al. 2008 ^11^ provided support for this hypothesis by showing that fetal inactivating heterozygous mutations in the glucokinase (*GCK*) gene, known to reduce fetal insulin secretion, reduces placental weight.

It is well established that the placenta plays a crucial role in the development of preeclampsia ^3,28^, and our results suggest that greater placental growth is on the causal pathway to disease. Dahlstrøm et al. 2008 et al ^29^ showed that both low and high placental weights were associated with increased risk of preeclampsia. The association of small placentas with preeclampsia was stronger in individuals diagnosed before 37 weeks of gestation. Those findings are in line with early-onset (diagnosis before 34 weeks) or pre-term (diagnosis before 37 weeks) preeclampsia believed to arise from poor placentation early in pregnancy ^1^. Where possible, we restricted our analysis to term pregnancies only, to include mainly late-onset preeclampsia cases. As the time of diagnosis was not known, we acknowledge that the timing of birth does not precisely represent the time of preeclampsia diagnosis. While our observational multivariable regression results did not indicate a relationship between placental weight and preeclampsia, we showed a robust causal association between higher placental weight and preeclampsia using one- and two-sample MR, consistent with previously reported two-sample MR results ^10^. It is possible that there was no observable association between placental weight and preeclampsia in our study due to the impact of reverse causation. In contrast, genetic instruments reflect the placenta’s capacity for growth and are not subject to reverse causation. Our finding of higher placental weight being causally linked to increased preeclampsia risk is in line with the hypothesis that placental function declines as it outgrows uterine capacity ^12^. Late decline in placental function is a potential cause of term preeclampsia ^12,30^. Based on our results we hypothesise that such a decline is reached earlier in pregnancies where the mother has diabetes than in pregnancies without diabetes due to increased insulin-mediated placental growth.

We performed several sensitivity analyses for two-sample MR. A high Q-statistic was identified for the two-sample MR analysis of fasting glucose on risk of preeclampsia, indicating more variability amongst the SNP-specific causal effect estimates than expected by chance. SNPs affecting the outcome not via the exposure (pleiotropy) can be a source of this additional variability. Furthermore, an MR-Egger intercept significantly different from zero is an indication for unbalanced pleiotropy. Hence, it is possible that the two-sample MR estimates for this pathway are biased due to unbalanced pleiotropy, which violates the exclusion restriction assumption, or several other assumptions that are key for the two-sample approach to be valid, and not for one-sample approaches. These include a miss-specified 1^st^ stage model, and the same distribution of confounders in the exposure and outcome GWAS population. Performing a range of sensitivity analyses improve the confidence and robustness of our results and highlight the importance of triangulating the results from different methods, a strength of our study.

Our study has a number of limitations. First, placental weight measured at delivery, although readily available in large samples of genotyped individuals, is a limited indicator of placental growth during pregnancy. For example, it does not capture placental surface area or density, both of which are likely relevant to preeclampsia risk. More studies using other placental measures are therefore necessary to fully understand how properties of the placenta influence preeclampsia. Second, it is possible that the relationship between placental weight and risk of preeclampsia is non-linear. Given the current understanding of late-onset preeclampsia relating to the placenta going beyond its capacity, it is possible that an increase in placental weight for already heavy placentas has a bigger effect on preeclampsia risk than an increase in placental weight for a light placenta. Future work could include the investigation of non-linear effects. A third limitation of our study is the use of maternal fasting glucose SNPs for the one-sample and two-sample MR analyses that were identified in a GWAS of non-pregnant individuals. Although this provided the strongest collective genetic instrument for fasting glucose, we made the assumption that genetic effects are the same/similar during and outside of pregnancy. This assumption was supported by the fact that the GRS comprised of “all fasting glucose SNPs” associated with maternal fasting glucose levels at 28 weeks of gestation in the EFSOCH study. Our sensitivity analyses using SNPs identified in a GWAS of fasting glucose during pregnancy had the advantage of genetic instruments that had been identified in pregnant participants. However, the GWAS was smaller and identified a lower number of SNPs. Furthermore, we assumed that fasting glucose effects are similar in mothers of Chinese and European ancestry. As the outcome GWAS used for the two-sample MR analyses was performed in a European population, we are at risk of violating the assumption that the exposure and the outcome GWAS should arise from a homogenous sample ^31^. We showed that a GRS of the “during pregnancy (Chinese) SNPs” also associated with maternal fasting glucose levels in pregnancy in EFSOCH, albeit to a smaller extent compared to the “all fasting glucose SNP” GRS. However, seeing consistent results in our two-sample MR analyses provides us with confidence in the validity of our findings.

To conclude, our results provide evidence that greater placental growth is on the causal pathway between higher maternal fasting glucose levels in pregnancy and risk of preeclampsia. A better understanding of causal biological processes underlying the pathology of preeclampsia is important for the development of treatment and optimal care for pregnant women. If confirmed, our results support that measuring and optimising maternal glucose levels in pregnancies affected by diabetes will be an important tool in reducing preeclampsia risk and suggest that markers of placental growth could help with risk stratification in the future.

## Data Availability

All data produced in the present work are contained in the manuscript. Information on how to request access to individual level data used in this study are described in the manuscript

## Acknowledgements

We are extremely grateful to all the families who took part in the EFSOCH study, the midwives for their help in recruiting them, and the whole study team, which includes interviewers, computer and laboratory technicians, clerical workers, research scientists, volunteers, managers, receptionists and nurses.

We are extremely grateful to all the families who took part in the ALSPAC study, the midwives for their help in recruiting them, and the whole ALSPAC team, which includes data collection staff, data and administrations staff, technical managers and the technical staff with the Bristol Bioresource Laboratory, based within the University of Bristol The authors would like to acknowledge the use of the University of Exeter High Performance Computing (HPC) facility in carrying out this work.

## Data Availability

The EFSOCH data set can be requested for access by writing in the first instance to the EFSOCH data team via the Exeter Clinical Research Facility.

The informed consent obtained from ALSPAC (Avon Longitudinal Study of Parents and Children) participants does not allow the data to be made available through any third party maintained public repository. Supporting data are available from ALSPAC on request under the approved proposal number, B3392. Full instructions for applying for data access can be found here: http://www.bristol.ac.uk/alspac/researchers/access/. The ALSPAC study website contains details of all available data (http://www.bristol.ac.uk/alspac/researchers/our-data/). Please note that the study website contains details of all the data that is available through a fully searchable data dictionary and variable search tool” and reference the following webpage:http://www.bristol.ac.uk/alspac/researchers/our-data/

## Ethical approval

Ethical approval for the study was obtained from the ALSPAC Ethics and Law Committee and the Local Research Ethics Committees. Informed consent for the use of all data collected was obtained from participants following the recommendations of the ALSPAC Ethics and Law Committee at the time. Participants can contact the study team at any time to retrospectively withdraw consent for their data to be used. Study participation is voluntary and during all data collection sweeps, information was provided on the intended use of data.

## Authors’ contributions

A.J., R.M.F., R.N.B and J.B., contributed to the study design. A.J. performed all analyses in this study, created figures and tables and drafted the manuscript. Data interpretation and statistical analysis were aided by R.N.B, J.B. and K.T.A.. Biological and clinical interpretation were supported by R.M.F. and A.E.H.. All authors reviewed and edited previous versions of the manuscript. All authors read and approved the final manuscript.

## Abbreviations

GWAS: Genome-wide association study
MR: Mendelian randomization
SD: Standard deviation
GRS: Genetic risk score
CI: Confidence interval

## References

1. Magee LA, Nicolaides KH, von Dadelszen P. Preeclampsia. Longo DL, editor. New England Journal of Medicine. 2022 May 12;386(19):1817–32.

2. Yang Y, Wu N. Gestational Diabetes Mellitus and Preeclampsia: Correlation and Influencing Factors. Front Cardiovasc Med. 2022;9.

3. Dimitriadis E, Rolnik DL, Zhou W, Estrada-Gutierrez G, Koga K, Francisco RPV, et al. Preeclampsia. Nat Rev Dis Primers. 2023;9(1).

4. Magee LA, Wright D, Syngelaki A, Von Dadelszen P, Akolekar R, Wright A, et al. Preeclampsia Prevention by Timed Birth at Term. Hypertension. 2023;80(5).

5. Weissgerber TL, Mudd LM. Preeclampsia and Diabetes. Curr Diab Rep. 2015;15(3).

6. Catalano PM, McIntyre HD, Cruickshank JK, McCance DR, Dyer AR, Metzger BE, et al. The hyperglycemia and adverse pregnancy outcome study: Associations of GDM and obesity with pregnancy outcomes. Diabetes Care. 2012;35(4).

7. The HAPO Study Cooperative Research Group. Hyperglycemia and Adverse Pregnancy Outcomes. New England Journal of Medicine. 2008;358(19).

8. Hufnagel A, Dearden L, Fernandez-Twinn DS, Ozanne SE. Programming of cardiometabolic health: the role of maternal and fetal hyperinsulinaemia. Journal of Endocrinology. 2022;253(2).

9. Hughes AE, De Franco E, Freathy RM, Flanagan SE, Hattersley AT. Monogenic disease analysis establishes that fetal insulin accounts for half of human fetal growth. Journal of Clinical Investigation. 2023;133(6).

10. Beaumont RN, Flatley C, Vaudel M, Wu X, Chen J, Moen GH, et al. Genome-wide association study of placental weight identifies distinct and shared genetic influences between placental and fetal growth. Nat Genet. 2023;55(11).

11. Shields BM, Spyer G, Slingerland AS, Knight BA, Ellard S, Clark PM, et al. Mutations in the Glucokinase Gene of the Fetus Result in Reduced Placental Weight. Diabetes Care. 2008;31(4).

12. Redman CWG, Staff AC, Roberts JM. Syncytiotrophoblast stress in preeclampsia: the convergence point for multiple pathways. Am J Obstet Gynecol. 2022;226(2).

13. Laine K, Murzakanova G, Sole KB, Pay AD, Heradstveit S, Raïsänen S. Prevalence and risk of pre-eclampsia and gestational hypertension in twin pregnancies: A population-based register study. BMJ Open. 2019;9(7).

14. Lawlor DA, Harbord RM, Sterne JAC, Timpson N, Davey Smith G. Mendelian randomization: Using genes as instruments for making causal inferences in epidemiology. Stat Med. 2008;27(8):1133–63.

15. Warrington NM, Beaumont RN, Horikoshi M, Day FR, Helgeland ø, Laurin C, et al. Maternal and fetal genetic effects on birth weight and their relevance to cardio-metabolic risk factors. Nat Genet. 2019;51(5):804–14.

16. Hemani G, Zheng J, Elsworth B, Wade KH, Haberland V, Baird D, et al. The MR-base platform supports systematic causal inference across the human phenome. Elife. 2018;7.

17. Sobel ME. Asymptotic Confidence Intervals for Indirect Effects in Structural Equation Models. Sociol Methodol. 1982;13:290.

18. Grotzinger AD, Fuente J de la, Privé F, Nivard MG, Tucker-Drob EM. Pervasive Downward Bias in Estimates of Liability-Scale Heritability in Genome-wide Association Study Meta-analysis: A Simple Solution. Biol Psychiatry. 2023;93(1).

19. Bowden J, Davey Smith G, Burgess S. Mendelian randomization with invalid instruments: Effect estimation and bias detection through Egger regression. Int J Epidemiol. 2015;44(2).

20. Dudbridge F, Voller B, Woodward RM, Frayling T, Pilling LC, Bowden J. Getting to GRIPS with MR-Egger: modelling directional pleiotropy independently of allele coding. 2025 Jun 24;

21. Bowden J, Davey Smith G, Haycock PC, Burgess S. Consistent Estimation in Mendelian Randomization with Some Invalid Instruments Using a Weighted Median Estimator. Genet Epidemiol. 2016;40(4).

22. Hartwig FP, Smith GD, Bowden J. Robust inference in summary data Mendelian randomization via the zero modal pleiotropy assumption. Int J Epidemiol. 2017;46(6).

23. Bowden J, Spiller W, Del Greco FM, Sheehan N, Thompson J, Minelli C, et al. Improving the visualization, interpretation and analysis of two-sample summary data Mendelian randomization via the Radial plot and Radial regression. Int J Epidemiol. 2018;47(4).

24. Chen J, Spracklen CN, Marenne G, Varshney A, Corbin LJ, Luan J, et al. The trans-ancestral genomic architecture of glycemic traits. Nat Genet. 2021;53(6).

25. Gu Y, Zheng H, Wang P, Liu Y, Guo X, Wei Y, et al. Genetic architecture and risk prediction of gestational diabetes mellitus in Chinese pregnancies. Nat Commun. 2025 May 5;16(1):4178.

26. Scott RA, Lagou V, Welch RP, Wheeler E, Montasser ME, Luan J, et al. Large-scale association analyses identify new loci influencing glycemic traits and provide insight into the underlying biological pathways. Nat Genet. 2012;44(9).

27. Honigberg MC, Truong B, Khan RR, Xiao B, Bhatta L, Vy HMT, et al. Polygenic prediction of preeclampsia and gestational hypertension. Nat Med. 2023;29(6).

28. Roberts JM, Escudero C. The placenta in preeclampsia. Pregnancy Hypertens. 2012;2(2).

29. Dahlstrøm B, Romundstad P, øian P, Vatten LJ, Eskild A. Placenta weight in pre-eclampsia. Acta Obstet Gynecol Scand. 2008;87(6).

30. Redman CWG, Staff AC. Preeclampsia, biomarkers, syncytiotrophoblast stress, and placental capacity. Am J Obstet Gynecol. 2015;213(4).

31. Zhao Q, Wang J, Spiller W, Bowden J, Small DS. Two-sample instrumental variable analyses using heterogeneous samples. Statistical Science. 2019;34(2).

