## Supplementary material for "Placental growth plays a key role in the link between maternal glucose levels in pregnancy and risk of preeclampsia"

### Supporting information

#### Study details EFSOCH and ALSPAC

The **Exeter Family Study of Childhood Health (EFSOCH)** is a prospective cohort study, which recruited families from central Exeter, United Kingdom (UK) [1]. Recruitment was completed in 2004 with most of the data collection being completed by 2006. Additional data, which included information on preeclampsia, was extracted from the individuals' medical records in 2020. We only included babies with gestational duration ≥ 37 weeks. We removed pre-term births to include mainly late-onset preeclampsia cases, as it is more likely that mothers with early-onset preeclampsia deliver earlier. Information on time of diagnosis for preeclampsia was not available. After removing individuals who did not meet the study requirements (for example withdrawn consents or non-European mother or father) and twin and pre-term pregnancies the study sample size was 948. Maternal-fetal pair information with genotypes was available in 636 pregnancies.

Pregnant women resident in Avon, UK with expected dates of delivery between 1st April 1991 and 31st December 1992 were invited to take part in the **Avon Longitudinal Study of Parents and Children (ALSPAC)** [2–4]. There were 20,248 eligible pregnancies and the initial number of pregnancies enrolled was 14,541. Of the initial pregnancies, there was a total of 14,676 fetuses, resulting in 14,062 live births and 13,988 children who were alive at 1 year of age. We restricted our analysis to singleton and term born pregnancies with available placental weight measures. The study sample size was 5,214. For analyses including genetic information we used unrelated mothers of European genetic similarity and their offspring including their genotypes, and the sample reduced to 2,045.

#### Descriptive statistics EFSOCH and ALSPAC

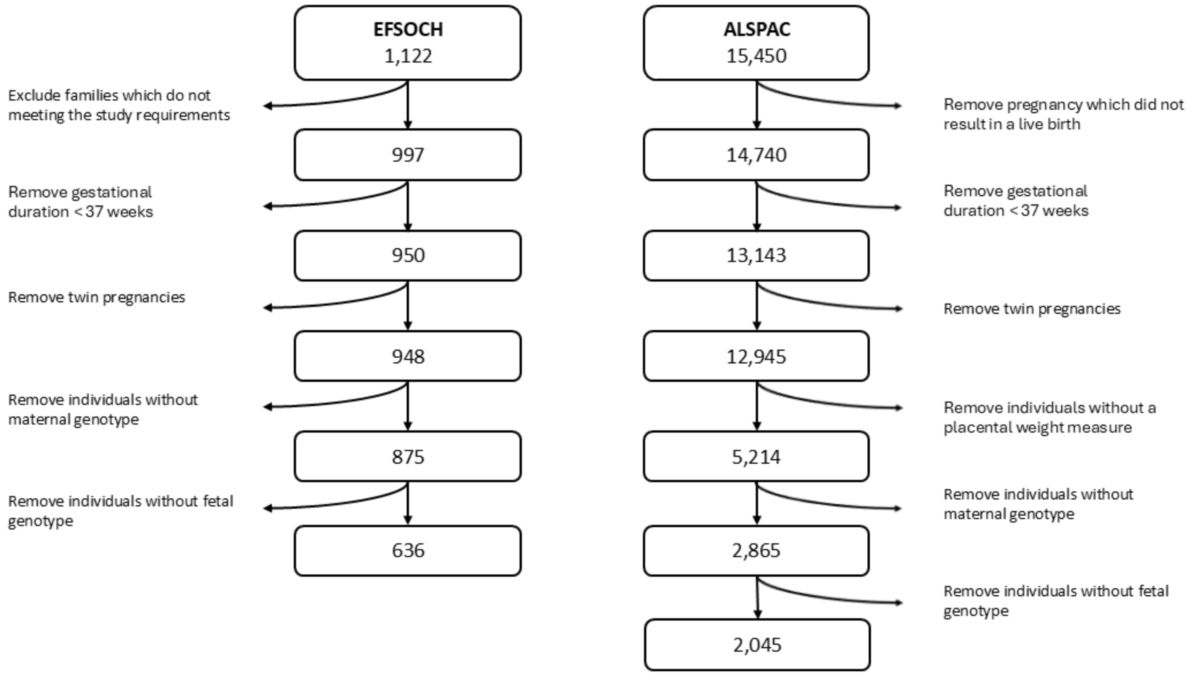


**Figure S1: Flowchart showing the sample size used for multivariable regression and one-sample MR analyses in EFSOCH and ALSPAC with indication of exclusions.**

**Table S1: Descriptive statistics in the EFSOCH dataset with and without genetic information**

| | **EFSOCH N = 948 (no genetic information included)** | | | **EFSOCH N = 636 (with fetal and maternal genetic information)** | | |
|---|---|---|---|---|---|---|
| | Mean/Cases | SD | Missing | Mean/Cases | SD | Missing |
| **Mothers fasting glucose at 28 weeks (mmol/l)** | 4.3 | 0.4 | 22 | 4.4 | 0.4 | 19 |
| **Placental weight (g)** | 635.8 | 134.1 | 112 | 635.9 | 134.6 | 76 |
| **Preeclampsia cases** | 25 (2.6 %) | NA | 28 | 14 (2.2 %) | NA | 15 |
| **Gestational duration (weeks)** | 40.1 | 1.2 | 0 | 40.1 | 1.2 | 0 |
| **Offspring sex (Male)** | 492 (51.9 %) | NA | 0 | 330 (51.9 %) | NA | 0 |
| **Birth weight (g)** | 3,503.9 | 473.7 | 0 | 3,518.5 | 474.4 | 0 |
| **Smokers (yes)** | 123 (13.0 %) | NA | 3 | 87 (13.7 %) | NA | 1 |
| **Maternal age** | 30.44 | 5.21 | 1 | 30.4 | 5.2 | 1 |
| **Parity (1 or more previous pregnancies)** | 513 (54.1 %) | NA | 0 | 359 (56.4 %) | NA | 0 |

**Table S2: Descriptive statistics in the ALSPAC dataset with and without genetic information.**

| | **ALSPAC N = 5,214 (no genetic information included)** | | | **ALSPAC N = 2,045 (with fetal and maternal genetic information)** | | |
|---|---|---|---|---|---|---|
| | Mean/Cases | SD | Missing | Mean/Cases | SD | Missing |
| **Mothers fasting glucose at 28 weeks (mmol/l)** | NA | NA | NA | NA | NA | NA |
| **Placental weight (g)** | 659.4 | 132.6 | 0 | 660.1 | 128 | 0 |
| **Preeclampsia cases** | 87 (1.7 %) | NA | 1 | 31 (1.5 %) | NA | 0 |
| **Gestational duration (weeks)** | 39.7 | 1.3 | 0 | 39.7 | 1.3 | 0 |
| **Offspring sex (Male)** | 2,664 (51.1 %) | NA | 0 | 998 (48.8 %) | NA | 0 |
| **Birth weight (g)** | 3,458.4 | 471.2 | 45 | 3,478.1 | 467.5 | 15 |
| **Smokers (yes)** | 1,212 (23.2 %) | NA | 274 | 373 (18.2 %) | NA | 51 |
| **Maternal age** | 27.9 | 4.9 | 0 | 28.5 | 4.6 | 0 |
| **Parity (1 or more previous pregnancies)** | 2,653 (50.9 %) | NA | 333 | 1,059 (51.8 %) | NA | 65 |

**Table S3: Descriptive statistics in the ALSPAC dataset with and without genetic information.**

| | Sample Size of GWAS | # Lead SNPs | Details |
|---|---|---|---|
| **All fasting glucose SNPs**<br>Chen et al. (2021) [5] | 281,416 | 105 | • Multi-ancestry GWAS<br>• SNPs that were significant in the analysis of European ancestry participants and added signals from the trans-ancestry analysis only if no variant in linkage disequilibrium (LD) already existed<br>• Fasting glucose is adjusted for BMI |
| **During pregnancy SNPs (Chinese)**<br>Gu et al. (2025) [6] | 85,086 | 42 | • Pregnant women of Chinese descent<br>• Fasting glucose in pregnancy between 24 and 28 weeks of gestation<br>• Fasting glucose is adjusted for BMI<br>• Effect estimates transformed to reflect increase/decrease in mmol/l (1SD = 0.42 mmol/l) |
| **Placental weight SNPs**<br>Beaumont et al. (2023) [7] | 65,405 fetal, 61,228 maternal | 37 fetal, 4 maternal | • GWAS performed in maternal and fetal genome<br>• Adjusted for the correlation between the maternal and fetal SNP effects using the weighted linear model adjustment approach [8] |
| **Preeclampsia SNPs**<br>Steinthorsdottir et al. (2020) [9]<br>Honigberg et al. (2023) [10] | 377,975 fetal, 723,181 maternal | Outcome only | • Analysis with fetal genome: 4,630 cases and 373,345 controls<br>• Analysis with maternal genome: 20,064 cases and 703,117 controls<br>• Adjusted for the correlation between the maternal and fetal SNP effects using the weighted linear model adjustment approach [8] |

**Additional observational estimates using EFSOCH and ALSPAC**

The univariate regression analysis estimates (unadjusted) for the observational associations between fasting glucose and placental weight, between placental weight and preeclampsia and between fasting glucose and preeclampsia are shown below. The HAPO study result has been previously published [11] and is presented for comparison. Additionally, we adjusted the analyses for offspring sex, maternal age, parity and smoking status during pregnancy. The regression estimates are shown in table S5.

**Table S4: Effect sizes for univariate regression analysis of the listed exposure on the listed outcomes in different individual level datasets.**

| Dataset | Exposure | Outcome | Effect [95 % CI] | P-value | N ($N_{effective}$) |
|---|---|---|---|---|---|
| EFSOCH | Fasting glucose (mmol/l) | Placental weight (g) | 81.6 [57.7,105.6] | $4.4 \times 10^{-11}$ | 818 |
| EFSOCH | Placental weight (SD) | Preeclampsia (Odds ratio) | 0.8 [0.5, 1.3] | 0.4 | 815 (89) |
| ALSPAC | Placental weight (SD) | Preeclampsia (Odds ratio) | 1 [0.8, 1.2] | 0.8 | 5,213 (342) |
| EFSOCH | Fasting glucose (mmol/l) | Preeclampsia (Odds ratio) | 4.1 [1.7, 9.4] | $1.1 \times 10^{-3}$ | 899 (97) |
| HAPO | Fasting glucose (mmol/l) | Preeclampsia (Odds ratio) | 2.6 [2.27,3.03] | $1.7 \times 10^{-38}$ | 25,505 (4,269) |

**Table S5: Effect sizes for regression analysis of the listed exposure on the listed outcomes adjusted for offspring sex, parity, maternal age and smoking status of the mother in EFSOCH and ALSPAC.**

| Dataset | Exposure | Outcome | Effect [95 % CI] | P-value | N ($N_{effective}$) |
|---|---|---|---|---|---|
| EFSOCH | Fasting glucose (mmol/l) | Placental weight (g) | 77.4 [52.9, 101.8] | $9.2 \times 10^{-10}$ | 816 |
| EFSOCH | Placental weight (SD) | Preeclampsia (Odds ratio) | 0.8 [0.5,1.2] | 0.3 | 813 (89) |
| ALSPAC | Placental weight (SD) | Preeclampsia (Odds ratio) | 1 [0.8, 1.2] | 0.9 | 4,870 (311) |
| EFSOCH | Fasting glucose (mmol/l) | Preeclampsia (Odds ratio) | 3.7 [1.6, 8.6] | $2.4 \times 10^{-3}$ | 896 (97) |

**Mendelian randomization (MR)**

MR builds on the concept of random inheritance of germline variants at conception. This random assortment, similar to a randomised clinical trial, makes this method less susceptible to confounding. Reverse causation can also be avoided as the genetic variants are usually not affected by disease state. [12]

There are three core assumptions that need to hold for a genetic variant to be a valid instrumental variable capable of furnishing accurate and unbiased causal estimates [13,14]:

1. The genetic variant needs to be strongly associated with the exposure of interest.
2. There are no confounders of the association between the genetic variant and the outcome.
3. The genetic variant is not associated with the outcome through any other pathways than via the exposure.

The first assumption is easy to verify, whereas the 2nd and 3rd assumptions are not; their plausibility must be carefully considered and investigated for the analysis at hand.

**Fasting glucose GRS and its association with maternal fasting glucose measured at 28 weeks of gestation in EFSOCH**

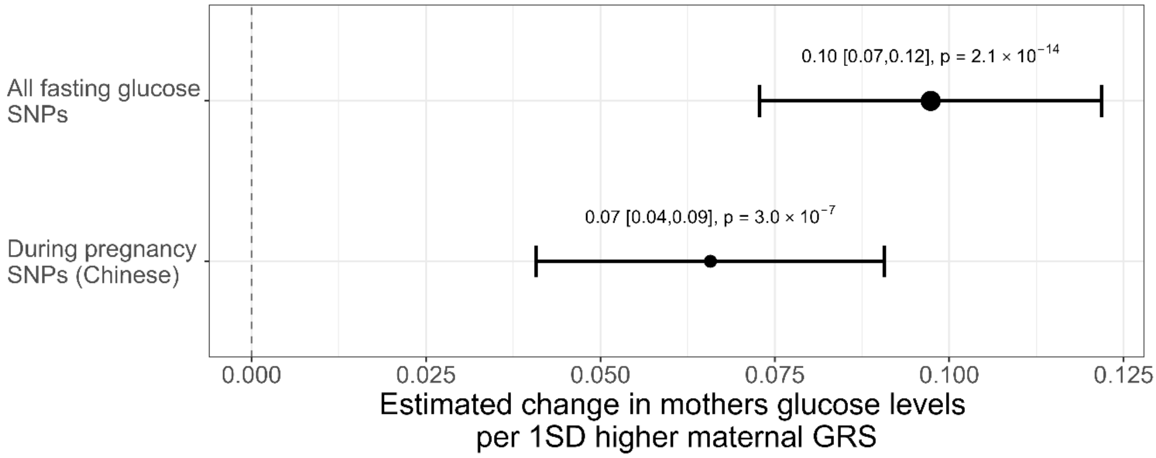


**Figure S2: Effect of maternal GRS, calculated with "all fasting glucose SNPs" and "during pregnancy SNPs (Chinese)", on maternal fasting glucose measured during pregnancy in EFSOCH.** 95% CI shown in squared brackets.

**Effective sample size**

The formula for the effective sample size is taken from Grotzinger et al. 2023 [15]

$$N_{effective} = 4 \cdot K \cdot (1 - K) \cdot (N_{cases} + N_{controls}),$$

with $K = \frac{N_{cases}}{N_{cases}+N_{controls}}.$

**Example of the two-stage least square (2SLS) regression model**

Equations 1 and 2 show the 2SLS model for the relationship between fasting glucose (FG) and placental weight (PW). Here the subscript $F$ and $M$ refer to fetal and maternal respectively. Variable $B$ reflects the genotyping batch variable. For analysis in ALPSAC $\alpha_3$ and $\beta_3$ are zero. $\widehat{FG}$ is the genetically predicted exposure obtained from Equation 1. The causal effect of fasting glucose on placental weight is estimated through $\beta_1$.

$$FG|GRS_M, Z = \alpha_0 + \alpha_1 GRS_M + \alpha_2 GRS_F + \alpha_3 B + \alpha_4 PC1 + \cdots + \alpha_{13}\, PC10 + \varepsilon_{FG} \tag{1}$$

$$PW|\widehat{FG}, Z = \beta_0 + \beta_1 \widehat{FG} + \beta_2 GRS_F + \beta_3\, B + \beta_4\, PC1 + \ldots + \beta_{13}\, PC10 + \epsilon_{PW} \tag{2}$$

**Delta method**

We derived the standard error for the two-step MR using the delta method as following:

$$\hat{\sigma}_{two-step} = \sqrt{\hat{\beta}^2_{FG,PW} \cdot \hat{\sigma}^2_{PW,PE} + \hat{\beta}^2_{PW,PE} \cdot \hat{\sigma}^2_{FG,PW}}\,,$$

where the subscripts FG, PW and PE refer to maternal fasting glucose, placental weight and preeclampsia respectively and the formula implicitly assumes that $\text{cov}(\hat{\beta}^2_{FG,PW}, \hat{\beta}^2_{PW,PE}) = 0$. The standard error, of the estimate referred to in the subscript, is denoted as $\sigma$. The estimates from the inverse variance weighted two-sample MR analyses are represented by $\beta$. The subscripts highlight from which analysis the effect sizes and the standard error, here the first listed abbreviation is the exposure and the second listed the outcome.

#### Number of SNPs and measures of instrument strength, heterogeneity and pleiotropy for each two-sample MR analysis (before outlier removal with MR-Radial)

**Table S6: Number of SNPs used for each two-sample MR analysis (before outlier removal) and measure of instrument strength (F-statistic) as well as investigations of heterogeneity (Q-statistic) and potential pleiotropy (MR-Egger intercept).** SNPs were removed and could not be included in the analysis if they were not available in the outcome GWAS data.
#: Number

| Exposure | Outcome | # Exposure SNPs | # SNPs used | SNPs removed | F-statistic | Q-Statistic (P-value) | MR-Egger intercept (P-value) |
|---|---|---|---|---|---|---|---|
| All fasting glucose SNPs | Placental weight (WLM maternal) | 105 | 99 | rs141107738, rs183381538, rs35188816, rs190355720, rs10717442, rs143960646 | 94.18 | 160.8 (0.00007) | 0.001 (0.65) |
| During pregnancy SNPs (Chinese) | Placental weight (WLM maternal | 42 | 39 | rs59789496, rs147272468 rs61020468 | 107.93 | 56.75 (0.025) | -0.0008 (0.8) |
| Placental weight (wlm fetal) | Preeclampsia (WLM fetal) | 37 | 34 | rs72804545, rs541641049, rs138715366 | 24.31 | 56.4 (0.007) | -0.01 (0.68) |
| Placental weight (wlm maternal) | Preeclampsia (WLM maternal) | 4 | 3 | rs72804545 | 28.97 | 0.36 (0.83) | 0.045 (0.69) |
| All fasting glucose SNPs | Preeclampsia (WLM maternal) | 105 | 97 | rs141107738, rs183381538, rs35188816, rs190355720, rs10717442, rs143960646, rs507666, rs189651013 | 95.25 | 150.9 (0.0003) | 0.023 (0.01) |
| During pregnancy SNPs (Chinese) | Preeclampsia (WLM maternal) | 42 | 38 | rs59789496, rs147272468, rs61020468, rs16922302 | 106.88 | 55.9 (0.02) | 0.001 (0.92) |

#### Comparing two-sample MR results before and after outlier removal

**Table S7: Number of SNPs used before and after outlier removal. Measure of instrument strength (F-statistic) as well as investigations of heterogeneity (Q-statistic) and potential pleiotropy (MR-Egger intercept) are shown after outlier removal using MR-Radial.** #: Number

| Exposure | Outcome | # SNPs before outlier removal | # SNPs used after outlier removal | F-statistic | Q-Statistic (P-value) | MR-Egger intercept (P-value) |
|---|---|---|---|---|---|---|
| All fasting glucose SNPs | Placental weight (WLM maternal) | 99 | 88 | 94.46 | 74.87 (0.82) | 0.0004 (0.8) |
| During pregnancy SNPs (Chinese) | Placental weight (WLM maternal | 39 | 35 | 104.8 | 36.22 (0.37) | -0.001 (0.64 |
| Placental weight (wlm fetal) | Preeclampsia (WLM fetal) | 34 | 29 | 24.25 | 24.94 (0.63) | -0.008 (0.74) |
| All fasting glucose SNPs | Preeclampsia (WLM maternal) | 97 | 85 | 87.91 | 74.71 (0.76) | 0.011 (0.11) |
| During pregnancy SNPs (Chinese) | Preeclampsia (WLM maternal) | 38 | 35 | 109.03 | 29.73 (0.68) | 0.003 (0.78) |

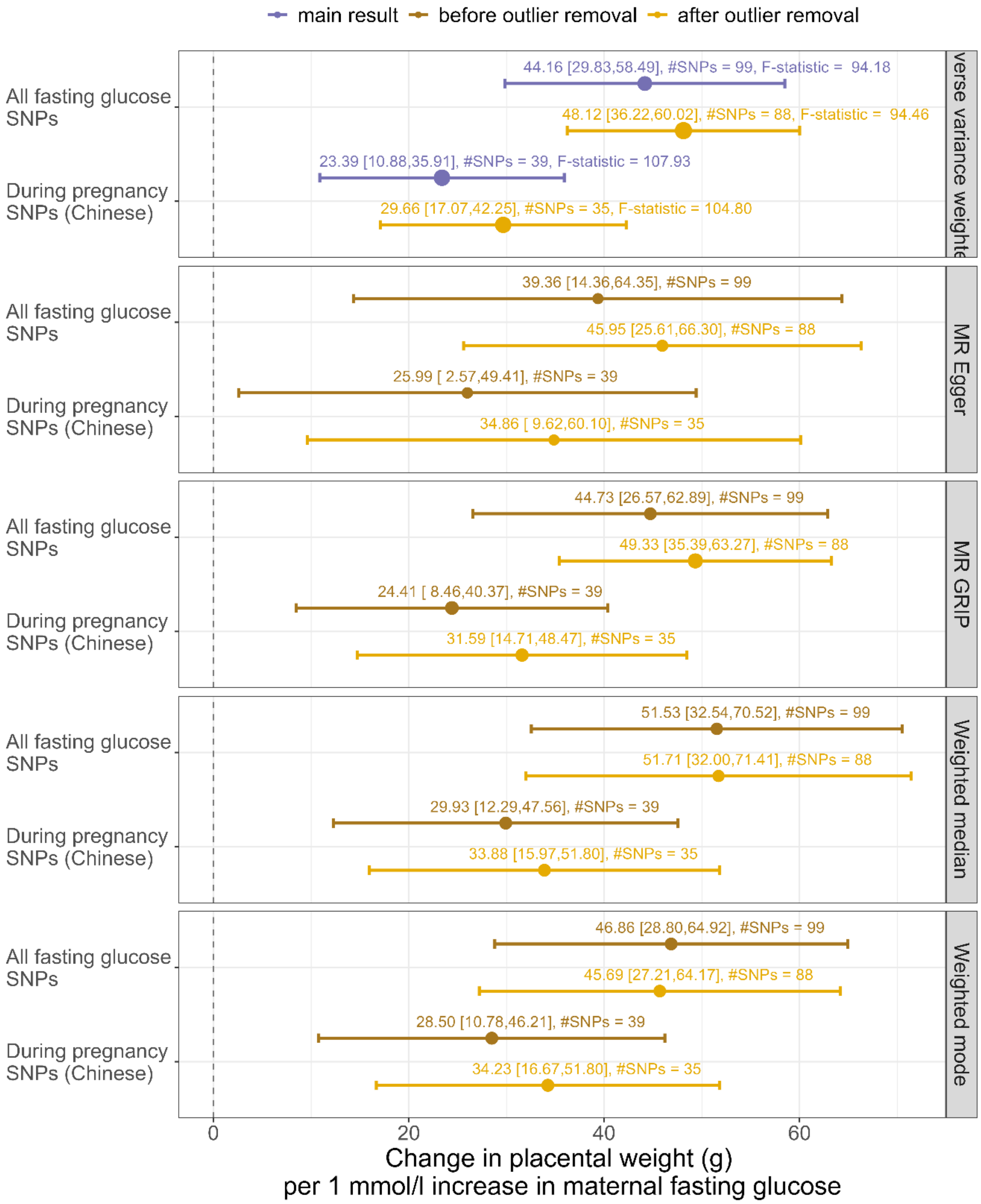


**Figure S3: Results from different two-sample MR analyses showing the effect of maternal fasting glucose on placental weight.** Results are shown before and after outlier removal (using MR-Radial). The result shown in the main paper (before outlier removal) is highlighted in purple. #SNPs represents the number of SNPs used for each analysis. 95% CI shown in squared brackets.

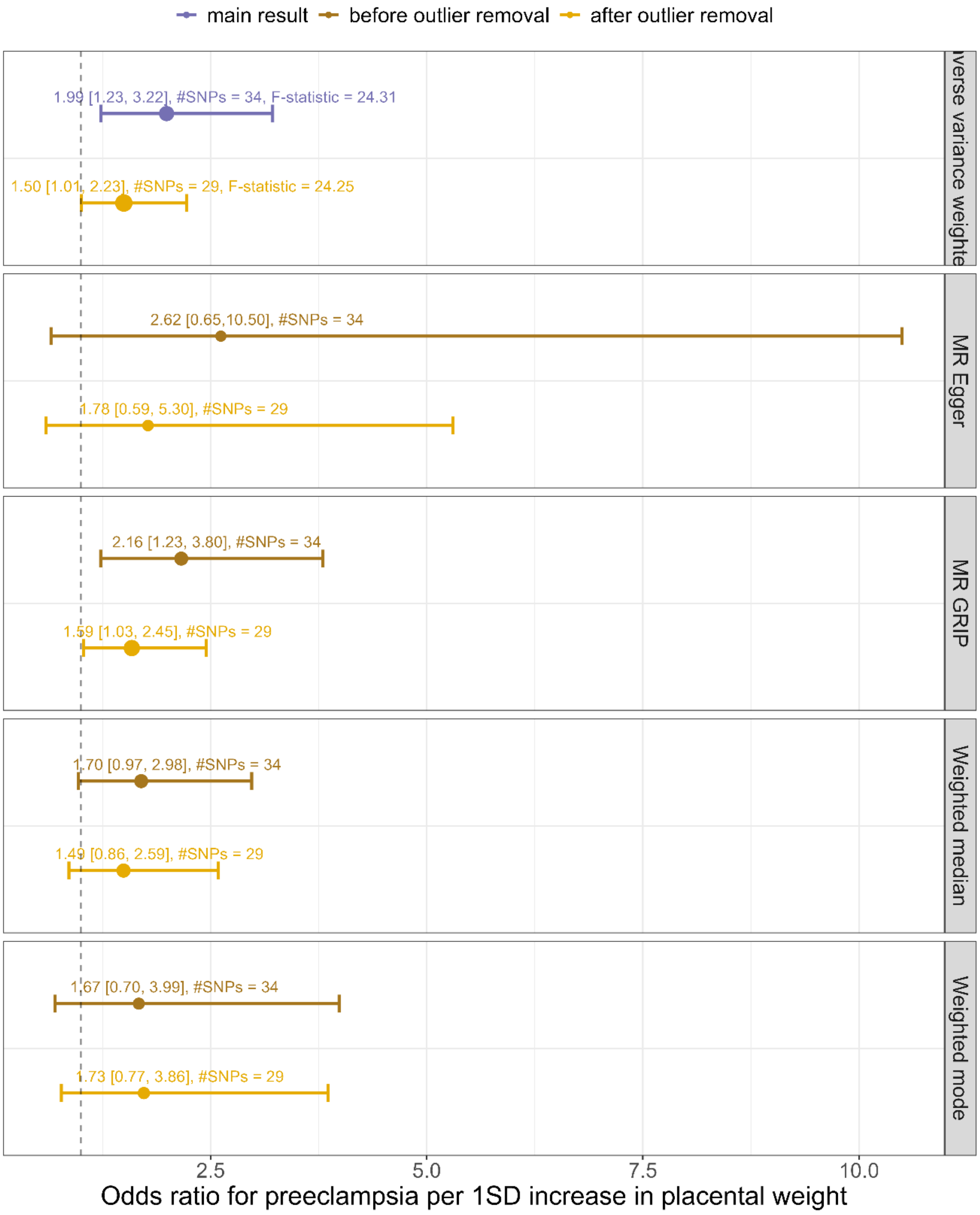


**Figure S4: Results from different two-sample MR analyses showing the effect of placental weight on risk of preeclampsia.** Results are shown before and after outlier removal (using MR-Radial). The result shown in the main paper (before outlier removal) is highlighted in purple. #SNPs represents the number of SNPs used for each analysis. 95% CI shown in squared brackets.

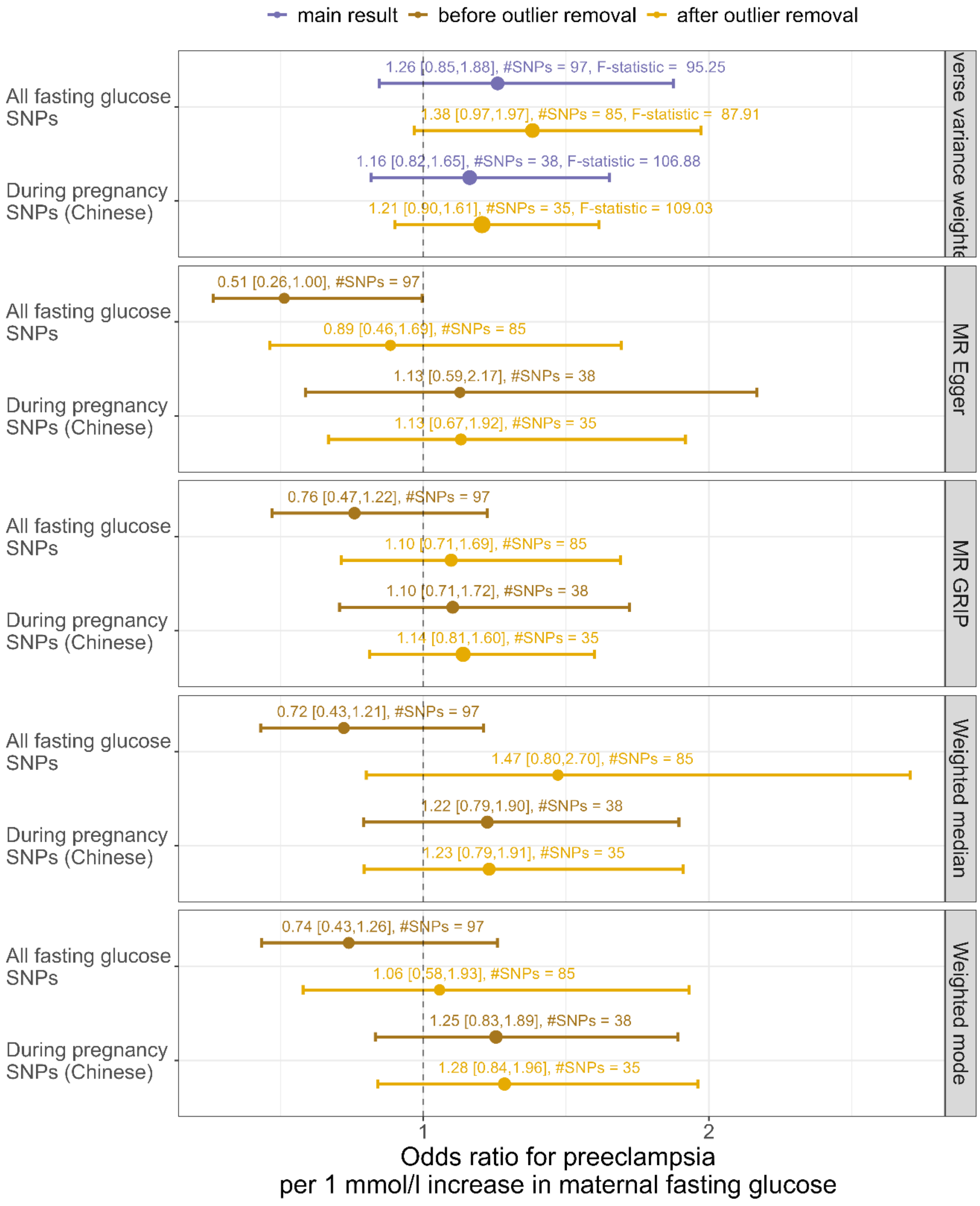


**Figure S5: Results from different two-sample MR analyses showing the effect of maternal fasting glucose on risk of preeclampsia.** Results are shown before and after outlier removal (using MR-Radial). The result shown in the main paper (before outlier removal) is highlighted in purple. #SNPs represents the number of SNPs used for each analysis. 95% CI shown in squared brackets.

1. Knight B, Shields BM, Hattersley AT. The Exeter Family Study of Childhood Health (EFSOCH): Study protocol and methodology. Paediatr Perinat Epidemiol. 2006;20(2).

2. Fraser A, Macdonald-wallis C, Tilling K, Boyd A, Golding J, Davey Smith G, et al. Cohort Profile: The Avon Longitudinal Study of Parents and Children: ALSPAC mothers cohort. Int J Epidemiol [Internet]. 2013 Feb 1 [cited 2022 Jul 14];42(1):97–110. Available from: https://academic.oup.com/ije/article/42/1/97/694445

3. Boyd A, Golding J, Macleod J, Lawlor DA, Fraser A, Henderson J, et al. Cohort Profile: The 'Children of the 90s'—the index offspring of the Avon Longitudinal Study of Parents and Children. Int J Epidemiol [Internet]. 2013 Feb [cited 2022 Jul 14];42(1):111. Available from: /pmc/articles/PMC3600618/

4. Northstone K, Ben Shlomo Y, Teyhan A, Hill A, Groom A, Mumme M, et al. The Avon Longitudinal Study of Parents and children ALSPAC G0 Partners: A cohort profile. [version 2; peer review: 1 approved]Wellcome Open Research. 2023;8.

5. Chen J, Spracklen CN, Marenne G, Varshney A, Corbin LJ, Luan J, et al. The trans-ancestral genomic architecture of glycemic traits. Nat Genet. 2021;53(6).

6. Gu Y, Zheng H, Wang P, Liu Y, Guo X, Wei Y, et al. Genetic architecture and risk prediction of gestational diabetes mellitus in Chinese pregnancies. Nat Commun. 2025 May 5;16(1):4178.

7. Beaumont RN, Flatley C, Vaudel M, Wu X, Chen J, Moen GH, et al. Genome-wide association study of placental weight identifies distinct and shared genetic influences between placental and fetal growth. Nat Genet. 2023;55(11).

8. Warrington NM, Beaumont RN, Horikoshi M, Day FR, Helgeland Ø, Laurin C, et al. Maternal and fetal genetic effects on birth weight and their relevance to cardio-metabolic risk factors. Nat Genet. 2019;51(5):804–14.

9. Steinthorsdottir V, McGinnis R, Williams NO, Stefansdottir L, Thorleifsson G, Shooter S, et al. Genetic predisposition to hypertension is associated with preeclampsia in European and Central Asian women. Nat Commun. 2020;11(1).

10. Honigberg MC, Truong B, Khan RR, Xiao B, Bhatta L, Vy HMT, et al. Polygenic prediction of preeclampsia and gestational hypertension. Nat Med. 2023;29(6).

11. The HAPO Study Cooperative Research Group. Hyperglycemia and Adverse Pregnancy Outcomes. New England Journal of Medicine. 2008;358(19).

12. Davey Smith G, Ebrahim S. 'Mendelian randomization': can genetic epidemiology contribute to understanding environmental determinants of disease? Int J Epidemiol. 2003 Feb 1;32(1):1–22.

13. Lawlor DA, Harbord RM, Sterne JAC, Timpson N, Davey Smith G. Mendelian randomization: Using genes as instruments for making causal inferences in epidemiology. Stat Med. 2008;27(8):1133–63.

14. Davies NM, Holmes M V., Davey Smith G. Reading Mendelian randomisation studies: A guide, glossary, and checklist for clinicians. BMJ (Online). 2018;362.

15. Grotzinger AD, Fuente J de la, Privé F, Nivard MG, Tucker-Drob EM. Pervasive Downward Bias in Estimates of Liability-Scale Heritability in Genome-wide Association Study Meta-analysis: A Simple Solution. Biol Psychiatry. 2023;93(1).
